# Disparities After Discharge: The Association of Limited English Proficiency and Post-Discharge Patient Reported Issues

**DOI:** 10.1101/2021.02.05.21251236

**Authors:** Lev Malevanchik, Margaret Wheeler, Kristin Gagliardi, Leah Karliner, Sachin J. Shah

## Abstract

**Background:** Patients with limited English proficiency (LEP) experience worse outcomes following hospital discharge. Care transition programs are common, yet little is known about the disparities in quality (process and outcome measures) experienced by patients with LEP.

**Methods:** We conducted a retrospective cohort study to determine the association of LEP and care transition quality at an urban, US academic hospital. We examined all adults discharge home from the hospital from May 2018 through April 2019. All patients received a multilingual, automated telephone call three days after discharge to assess and address patient-reported issues. We determined care transition quality using process measures (reach rate, time to resolve reported issue) and outcome measures (discharge instructions questions, difficulty obtaining prescriptions, medication concerns, follow up care questions, new or worsening symptoms, any other clinical issues). All results were adjusted for measured confounders; we used predicted probabilities and average marginal effect (AME) to describe associations.

**Results:** 13,860 patients were included in the study; 11% had LEP. The program reached most patients regardless of LEP status but was less likely to reach patients with LEP (AME 3.3%; 95% CI, 1.4% to 5.1%). After adjustment, patients with LEP reported high rates of all measured patient-reported outcomes: discharge instruction (AME 4.8%; 95% CI, 2.7% to 6.9%), obtaining prescriptions (AME 2.9%; 95% CI, 0.6% to 5.1%), medications concerns (AME 2.3%; 95% CI, 0.0% to 4.6%), follow up questions (AME 2.8%; 95% CI, 0.3% to 5.3%), new or worsening symptoms (AME 3.2%; 95% CI, 0.7% to 5.8%), and any other clinical issues (AME 3.6%; 95% CI, 1.1% to 6.1%). When issues were identified, the association between LEP and time to resolution of an issue was statistically, but not clinically, significant.

**Conclusion and Relevance:** Among patients with LEP, substantial disparities exist in patient-reported post-discharge outcome measures indicating a need for better discharge processes that focus on quality and health equity. Following discharge, relatively minor disparities were observed in the processes of a care transitions program that supports non-English languages.

## Introduction

Patients with limited English proficiency (LEP) commonly face barriers in communicating with their clinicians and understanding their treatment plans.^1^ These barriers are particularly problematic in the hospital, where patients with LEP are more likely to suffer adverse events, have higher mortality rates, and have higher readmission rates than English proficient (EP) patients.^2,3^ While prior studies have documented in-hospital adverse events, the experience of patients with LEP during and after hospital discharge is not well understood.

The transition from hospital to home is a vulnerable period for all patients,^4,5^ especially for those who have LEP.^6^ To identify and address issues that arise during this transition, hospitals have implemented programs to contact all recently discharged patients routinely. These programs have been shown to improve patient satisfaction and increase post-discharge medication adherence.^7^ To reach discharged patients efficiently, care transition programs have increasingly relied on technological solutions such as automated phone calls and text messages. Whether such tools effectively reach those with LEP is not known; however, evidence from prior technology implementation suggests that new technologies tend to widen healthcare disparities.^8,9^

Additionally, little is known about the issues and barriers that patients with LEP face after hospital discharge. Although one study found that patients with LEP have poor comprehension of hospital discharge instructions,^10^ other dimensions of the care transition such as worsening symptoms, difficulty obtaining prescriptions, or issues taking medications have not been described.

To address these two knowledge gaps in the care of patients with LEP, we examined data from a care transitions outreach program at a large academic medical center. Our first goal was to evaluate whether the program’s technology and processes reached patients with LEP and EP (English proficiency) equally. Our second goal was to measure the prevalence of post-discharge, patient-reported issues, and their association with English proficiency.

## Methods

### Design, Setting, Subjects

We performed a retrospective cohort study of patients 18 years or older discharged home from an academic medical center between May 1, 2018, and April 30, 2019. We included all patients who were discharged home from participating clinical services and were not part of a bundled payments program that coordinated post-discharge care separately. We excluded patients discharged with home hospice (1.2%), lacking a listed telephone number (0.4%), were admitted for less than one day (0.5%), were readmitted to the same hospital within 72 hours (1.8%), or were missing exposure covariate, or outcome data (2.8%). Additionally, we excluded 4.8% of discharges because medical records could not be matched to automated calling program data (**Appendix-1**). For patients with multiple admissions during the study period, we examined the first discharge.

### The Care Transitions Outreach Program

The study site implemented a hospital-wide care transitions outreach program in March 2017. The program’s goal was universal contact with all patients discharged from the hospital to identify and address care transition problems. In their discharge instructions (written in English), patients were notified that they would receive an automated call within 72 hours of discharge. A third party (CipherHealth Voice, New York, NY, USA) delivered a scripted, automated call to ask patients if they had questions or concerns in six post-discharge domains. The program called all patients regardless of their reported language preference. Respondents could choose to hear and respond to the questions in English, Spanish, or Cantonese, the three most common languages spoken by patients in the health system. The Appendix provides the complete text of the script (**Appendix-2**).

The automated calling program attempted to call patients up to five times per day for two consecutive days. If a patient failed to answer the automated call and met any of the following specific criteria—age >85 years, discharged home with home services, limited English proficiency, or part of the study site’s Medicare accountable care organization—a centralized care transition nurse would review the patient’s chart and call the patient manually if they had not already been contacted or seen by a clinician or their staff. If a patient did answer the automated call initially and identified an issue, the care transition nurse would call the patient manually to follow up. In either situation, the nurse would use a professional language interpreter if they were not fluent in the patient’s preferred language.

### Measures: Limited English proficiency

There is no gold standard for defining limited English proficiency. We classified patients as having limited English proficiency (LEP) if in the electronic medical record (EMR), their preferred language for healthcare was a language other than English and if they self-identified as needing an interpreter. This definition was validated through chart review in a prior study conducted at the same institution^3^. In that study, compared to the chart reviewers’ assessment of English proficiency, the definition had a positive predictive value of 100%; thus, any misclassification in this study would err on the side of categorizing patients with LEP as EP. Preferred language and need for an interpreter were obtained on registration and directly reflected the patients’ stated responses.

### Measures: Sociodemographics and clinical comorbidities

We obtained demographic and clinical data using structured elements from the EMR. Using administrative data we determined patients’ discharge diagnoses with International Classification of Diseases, Tenth Revision, Clinical Modification (ICD-10-CM) diagnosis codes; we used these codes to calculate the Elixhauser score for each patient.^11^

### Outcomes: Reach rate

We classified “patient reach” into one of four mutually exclusive, hierarchical categories. If a patient was able to answer the automated call and at least one clinical question, we classified them as having “answered an automated call.” If a patient did not answer the automated call but was contacted a clinician or their staff, we classified them as having been “contacted by a clinician.” For patients who neither answered the automated calls nor were reached by clinicians, the care transition nurses attempted to reach the patients who met the criteria listed above with up to three manual phone calls. If the nurses successfully reached a patient by manual call, we classified them as having been “reached by a manual call.” We classified patients as “not reached” if the automated calls, clinicians, and the care transitions nurses were unable to reach them.

### Outcomes: Patient-reported, post-discharge issues

The care transitions outreach program asked patients about six post-discharge issues: questions about their discharge instructions, difficulty getting their prescriptions, medication concerns, questions about follow up care, new or worsening symptoms, or any other clinical issues not already addressed (specific questions listed in **Appendix-2**).

In sensitivity analyses, we further characterized the severity of issues in two ways. First, following an outreach call, care transition nurses noted the urgency of reported issues using templated documentation, and categorized these from least to most urgent as follows: the nurse counseled the patient on the phone, the nurse notified a clinician, the nurse requested a non-urgent action by a clinician, or the nurse needed urgent action by the clinician within 24 hours. Second, when an issue was identified, we determined how often nurses needed to involve other healthcare professionals such as nonclinical health staff (e.g., social workers, case managers), clinicians, or pharmacists in order to resolve the issue.

### Analysis

To compare baseline variables, we used the chi-square test for categorical variables. For continuous variables, we used the t-test for normally distributed measures and the Wilcoxon rank sum test for nonnormal measures. Because successful outreach was categorized hierarchically, we fit an ordered logistic regression to determine the association with LEP and successful outreach. We determined the association between LEP and each post-discharge issue by fitting a separate generalized estimating equation with a log link and Poisson distribution^12^ for each patient-reported issue. For each model, patients were included if they responded to the question corresponding to the post-discharge issue (see **Appendix-2**). To determine the association between LEP and time to resolution of post-discharge issues, we fit a Cox proportional hazard model; this model was limited to patients who reported one or more issues. We verified the proportional hazards assumption by testing the interaction of LEP and time. All models were adjusted for the following confounders: age, race, ethnicity, marital status, insurance, discharging service, Elixhauser score, discharge with home services, and length of stay. We report the regression results as the predicted population rates and average marginal effects (AME) along with the 95% confidence intervals^13^. We completed analyses in SAS 9.4 (Cary, NC) and STATA 15.1 (College Station, TX). The UCSF Committee on Human Research approved this study. The code used to generate the cohort and perform the analyses can be found at https://github.com/sachinjshah.

## Results

### Participant Characteristics

During the one-year study period, 13,860 unique patients were discharged home from inpatient hospitalizations, and 1,566 (11%) had LEP (**Table 1**). The most common languages spoken by patients with LEP were Spanish (42%) and Cantonese (23%). Compared with English speakers, patients with LEP were more likely to be older (median 65y vs. 57y), Hispanic/Latinx (41% vs. 13%), and Asian (44% vs. 12%). Patients with LEP were more likely to be insured through Medicaid (37% vs. 21%) or Medicare (49% vs. 37%). Finally, patients with LEP were more likely to be discharged home with services (24% vs. 16%).

**Table 1:** Patient characteristics of discharged patients by English proficiency.

|  | Limited<br>English<br>proficiency<br>(n=1566) | English<br>proficient<br>(n=12294) | p value |
| --- | --- | --- | --- |
| Age, median years [IQR] | 65 [52, 77] | 57 [42, 67] | <0.001 |
| Male (%) | 789 (50) | 6467 (53) | 0.098 |
| Race (%) |  |  | <0.001 |
| Asian | 682 (44) | 1444 (12) |  |
| Black or African American | 9 (1) | 1145 (9) |  |
| Other | 640 (41) | 2017 (16) |  |
| White or Caucasian | 235 (15) | 7688 (63) |  |
| Hispanic ethnicity (%) | 635 (41) | 1657 (13) | <0.001 |
| Marital status (%) |  |  | <0.001 |
| Divorced/Separate | 105 (7) | 979 (8) |  |
| Married/Partnered | 994 (63) | 6593 (54) |  |
| Single | 253 (16) | 4136 (34) |  |
| Widowed | 214 (14) | 586 (5) |  |
| Insurance status (%) |  |  | <0.001 |
| Commercial | 203 (13) | 5133 (42) |  |
| Medicaid | 576 (37) | 2535 (21) |  |
| Medicare | 763 (49) | 4504 (37) |  |
| Other | 24 (2) | 122 (1) |  |
| Clinical service (%) |  |  | <0.001 |
| General Surgery | 175 (11) | 1535 (12) |  |
| Hospital Medicine | 455 (29) | 2597 (21) |  |
| Neurosurgery | 126 (8) | 1628 (13) |  |
| Other | 810 (52) | 6534 (53) |  |
| Length of stay (median [IQR]) | 3.2 [2.0, 5.8] | 3.1 [1.5, 5.4] | <0.001 |
| ICU stay (%) | 196 (13) | 1644 (13) | 0.368 |
| Elixhauser score (mean (SD)) | 2.0 (1.8) | 2.3 (1.8) | <0.001 |
| Discharge home with services (%) | 372 (24) | 1970 (16) | <0.001 |

IQR - interquartile range; ICU - intensive care unit
Of the 1,566 patients with LEP, the most common reported preferred languages were Spanish (42%) Cantonese (23%), Russian (8%), and Mandarin (7%). All other languages each made up less than 4% of discharges for patients with LEP. All of the patients with LEP self-identified as needing an interpreter.

### Limited English Proficiency and Patient Outreach

The care transition program reached a majority of discharged patients, although there were some differences by English proficiency (**Table 2**, unadjusted results **Appendix-3**). Fewer patients with LEP answered automated calls compared with patients with EP (average marginal effect [AME] −4.3%; 95% CI −1.9% to −6.7%). When patients did not answer the automated call, chart review identified that patients with LEP were more likely to have been contacted by a clinician or their staff (AME 0.9%; 95% CI 0.4% to 1.3%). When the care transition nurses called manually, they were statistically more likely to reach patients with LEP than those with EP, although the difference was not clinically significant. Overall, while most patients were reached by some method regardless of language status, patients with LEP were more likely not to be reached (AME 3.3%; 95% CI 1.4% to 5.1%).

**Table 2:** Post-discharge program reach by English proficiency, adjusted probability and marginal effect.

| **Patient outreach** | **Limited English proficiency, predicted probability** | **English proficient, predicted probability** | **Average marginal effect of limited English proficiency on outcome (95% CI)** |
| --- | --- | --- | --- |
| Answered automated call | 76.7% | 81.0% | -4.3% (-1.9% to -6.7%) |
| Outreached completed by clinician | 5.8% | 4.9% | 0.9% (0.4% to 1.3%) |
| Reached by manual RN call | 1.4% | 1.2% | 0.2% (0.1% to 0.3%) |
| Not reached by any method | 16.2% | 12.9% | 3.3% (1.4 to 5.1%) |

Categories are mutually exclusive, hierarchical, and modeled using an ordered logistic model. Predicted probabilities from the adjusted model are presented. All models were adjusted for the following confounders: age, race, ethnicity, marital status, insurance, discharging service, Elixhauser score, discharge disposition, length of stay. Group p value <0.001. Unadjusted results can be found in Appendix 3.

### Limited English Proficiency and Patient Outcomes

Patients with LEP were more likely to report a problem with all measured post-discharge issues (**Figure 1**, unadjusted results **Appendix-3**). A greater number of patients with LEP had questions regarding information in their discharge instructions compared to patients with EP (AME 4.8%; 95% CI, 2.7% to 6.9%). More patients with LEP needed help to get their prescriptions filled (AME 2.9%; 95% CI, 0.6% to 5.1%) and had concerns about their medications (AME 2.3%; 95% CI, 0.0% to 4.6%). After discharge, patients with LEP were more likely to have questions about follow up care (AME 2.8%; 95% CI, 0.3% to 5.3%). They were also more likely to have new or worsening symptoms (AME 3.2%; 95% CI, 0.7% to 5.8%), and other clinical questions for the nurses (AME 3.6%; 95% CI; 1.1% to 6.1%). Although patients with LEP faced more post-discharge issues, there was no significant difference in issue severity, as defined by the need for assistance after the phone call (**Appendix-4**). Furthermore, when patients reported an issue and the care transition nurse escalated the concern, nurses involved nonclinical health staff, clinicians, and pharmacists at similar rates (**Appendix-5**).

**Figure 1:**
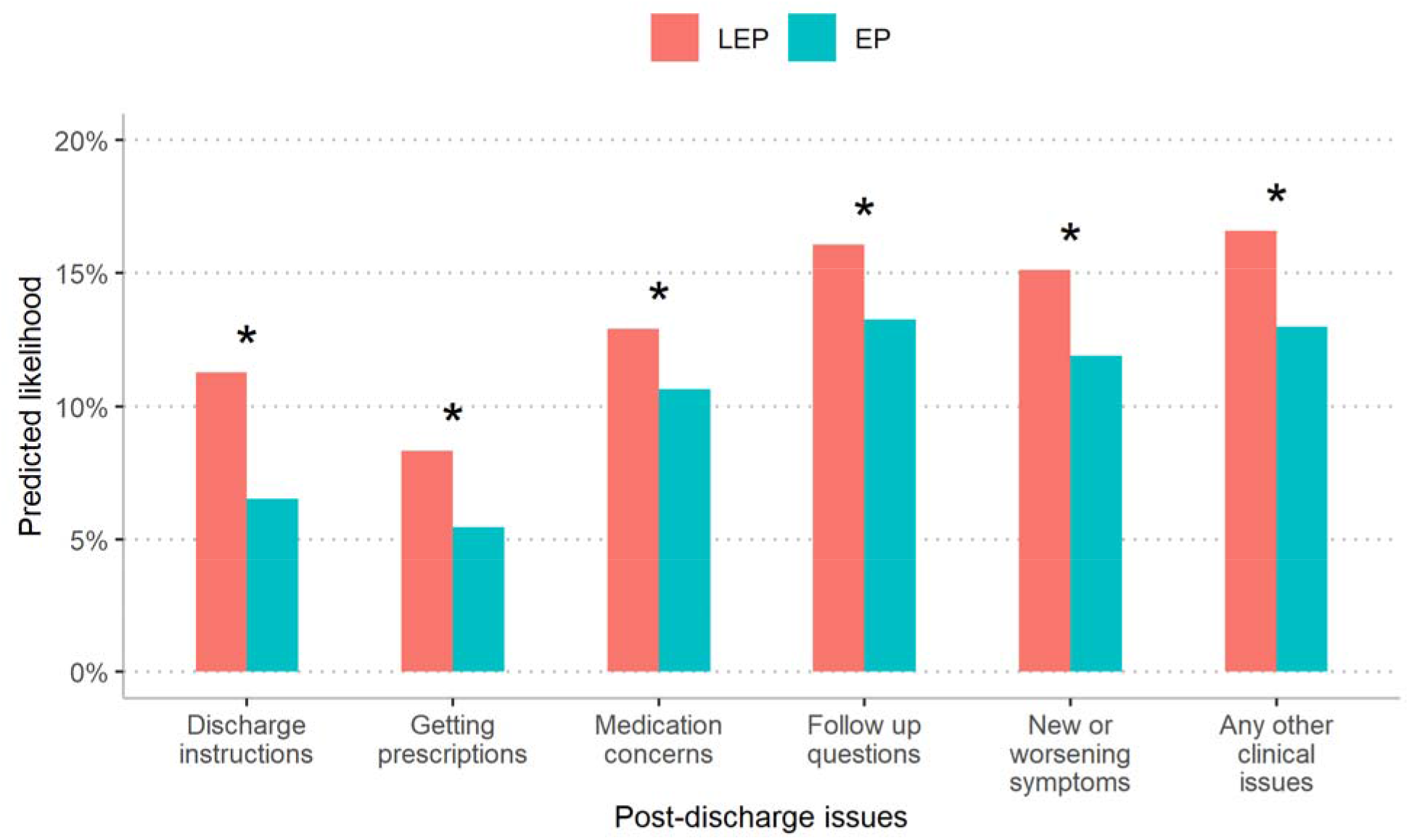
Patient-reported, post-discharge issues by English proficiency, adjusted. <u>Legend</u> LEP - limited English proficiency, EP - English proficient Predicted probabilities from the adjusted model are presented. Separate models were fit for each issue and included patients who answered the question: discharge instruction (n=10458), getting prescriptions (n=7849), medication concerns (n=10693), follow up questions (n=10554), new or worsening symptoms (n=11224), any other clinical issues (n=10170). Models were adjusted for the following confounders: age, race, ethnicity, marital status, insurance, discharging service, Elixhauser score, discharge disposition, length of stay. Asterisk denotes that the difference between groups is statistically significant with p < 0.05. Discharge instruction difference 4.8% (95% CI, 2.7% to 6.9%) p<0.001; Getting prescriptions difference 2.9% (95% CI, 0.6% to 5.1%) p=0.012; Medications concerns difference 2.3% (95% CI, 0.0% to 4.6%) p=0.0495; Follow up questions difference 2.8% (95% CI, 0.3% to 5.3%) p=0.027; New or worsening symptoms difference 3.2% (95% CI, 0.7% to 5.8%) p=0.012; Any other clinical issues difference 3.6% (95% CI, 1.1% to 6.1%) p=0.004. Unadjusted results can be found in Appendix 3.

Among the 3,895 patients who reported at least one post-discharge issue, having LEP was associated with a longer time to issue resolution (adjusted HR 0.84, 95% CI 0.75 to 0.93) (**Figure 2**, unadjusted results **Appendix-3**). That is, at any time point, patients with LEP were 84% as likely as EP patients to have had their issue resolved. While this difference was statistically significant, it did not translate to a clinically meaningful difference (predicted time to resolution, median 3.3 days vs. 3.1 days, p=0.001).

**Figure 2:**
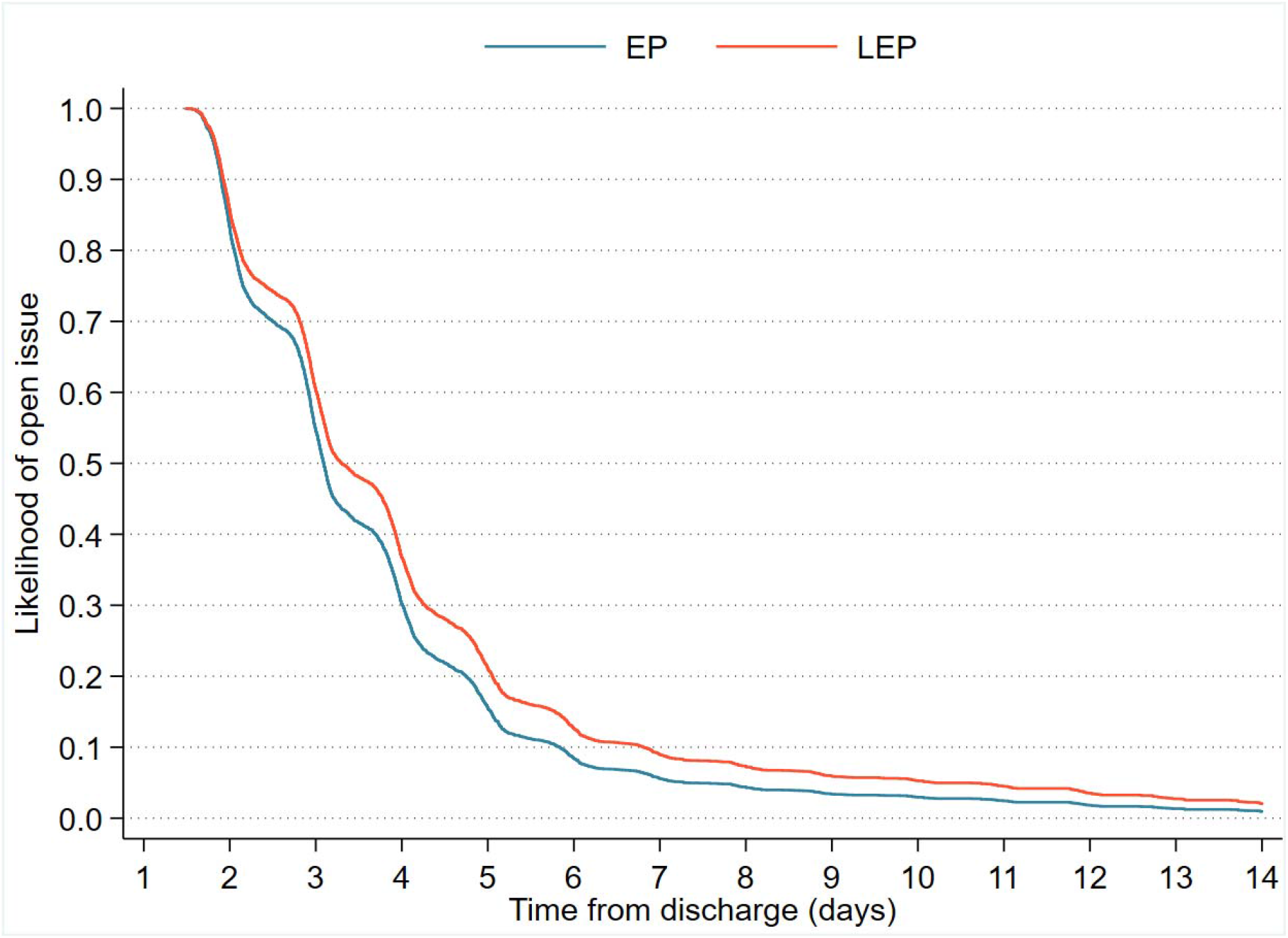
Time to issue resolution after discharge by English proficiency, adjusted. <u>Legend</u> LEP - limited English proficiency, EP - English proficient Plot depicts the likelihood of a patient’s reported issue being resolved at a given time post discharge by EP status. The likelihood is based on the Cox regression analysis and accounts for the following confounders: age, race, ethnicity, marital status, insurance, discharging service, Elixhauser score, discharge disposition, length of stay. The adjusted hazard of an open issue for patients with LEP is 0.84 (95%CI 0.75 to 0.93) relative to patients with EP. That is, at any time point, patients with LEP at 84% as likely as EP patients to have had their issue resolved. Predicted time to resolution, median 3.3 days for LEP, 3.1 days for EP (p=0.001). Unadjusted results can be found in Appendix 3.

## Discussion

Overall, the care transitions program reached a substantial proportion of patients discharged from our adult hospital, including both English and non-English speaking patients. The program’s robust efforts to reach patients with LEP, using multi-lingual automated scripts and prioritizing patients with LEP for manual outreach, enabled the program to reach a majority of patients with LEP. Despite these efforts, the program reached 3% fewer patients with LEP than those with EP.

Additionally, we found that limited English proficiency was strongly associated with higher rates of patient-reported, post-discharge issues. These associations were clinically significant and robust, persisting after accounting for confounders. Despite these disparities in reported post-discharge issues for patients with LEP, there were no differences in the urgency of reported issues or the need for assistance from other professionals. Additionally, the time needed to resolve patient-reported issues was similar for all patients.

The findings from this study confirm and expand on existing literature related to the communication barriers faced by hospitalized patients with LEP, which can manifest throughout the hospital course from admission to discharge. When patients with LEP are hospitalized, they receive adequate informed consent less than a third of the time, a statistic which is improved by easy access to professional interpreters.^14^ When critically ill, patients with LEP and their families receive less information and emotional support from their healthcare providers.^15^ During the hospital discharge process, few have a professional interpreter at the bedside to assist with discharge instructions,^6^ so after hospital discharge, it may not be surprising that patients with LEP report all post-discharge issues at higher rates. This study finds discharge disparities in multiple domains, from understanding instructions to follow up appointments to symptoms. Structural barriers, both during the hospital course and the care transition period, may explain the disparities patients with LEP face in outcomes like readmission and mortality.^16,17^

Clinicians and health systems can take steps to improve the discharge experience for patients with LEP. First and foremost, clinicians must use a professional interpreter such as a telephone, video, or in-person interpreter to communicate discharge instructions effectively to patients with LEP.^18^ Additionally, written translation technology can play an increasingly important role as technology improves. While the current implementation is imperfect, future iterations of artificial intelligence driven translation services may provide real-time, scalable text translation for written discharge instructions.^19^ It will be important to ensure that these translations are accurate in order to avoid adverse events post-discharge. One emerging program, Meds-to-Beds, relies on pharmacists to deliver medications and medication education to patients in the hospital before discharge, utilizing professional interpreters when needed, and has been shown to reduce readmission rates.^20^ Utilizing a program such as Meds-to-Beds along with appropriate written translations for patients to take home with them could lead to less miscommunication at the pharmacy when patients with LEP attempt to obtain their medications, a task often performed without an interpreter’s help. Finally, post-discharge population health programs are commonly used to identify issues that may otherwise go unaddressed. These programs rely on automated phone calls and text messages to scale outreach. To promote equity, population health programs should support non-English languages and examine their data to ensure they are reaching patients with LEP as we have done with the program reported here. These methods of improving care for patients with LEP are established but require health system leadership and buy-in, including financial resources.

Addressing disparities for patients with LEP during care transitions will require health systems to assess the underlying causes of the disparities, including communication barriers, access to professional medical interpreters, and systemic bias. However, there are already several available interventions available that can improve outcomes for patients with LEP. The first step in understanding barriers is to have robust data collection, including the appropriate classification of patients by English proficiency.^21,22^ Next, hospital systems must provide professional interpreters to all patients with LEP as mandated by the Civil Rights Act of 1964.^23,24^ Professional interpreters improve patients’ communication and understanding of their medical care.^14,25^ However, the provision of interpreter services does not guarantee adequate use; studies have documented that even when interpreters are available, they may not be routinely used.^26^ To assess interpreter use and support quality improvement programs to improve use, the electronic medical record must have a way for clinicians to easily document professional interpreter use during language discordant encounters.^21^ This critical intervention is only effective if it is utilized.

This study has several limitations inherent to the data available and the nature of the care transitions outreach program. This study was conducted at an urban academic medical center with a substantive and linguistically diverse LEP population, and the findings may not generalize to other settings. There is the potential for unmeasured confounding to influence our observed relationship between LEP and post-discharge issues, although we accounted for demographic variables, insurance status, patient complexity, and clinical service. Our criteria for defining LEP favored specificity over sensitivity; as such, there may be a small group of patients in the EP cohort who required language assistance during their hospital stay. This tradeoff biases the measured disparities towards the null; that is, the true disparity may be more substantial than what is measured in this study.

In this study, we demonstrated that a care transitions outreach program that supports non-English languages can reach a majority of patients with LEP. We also showed the importance of such an outreach program for patients with LEP as they experienced substantially more post-discharge issues than English speakers across many domains. Future work should focus on implementing and refining known solutions that improve the quality of hospital discharges and care transitions for patients with limited English proficiency.

## Data Availability

Researchers can contact the corresponding author to access to the data. Code used to generate the cohort and perform the analyses can be found on github (https://github.com/sachinjshah).

## Contributors

LM and SJS were responsible for the study concept and design. SJS obtained funding and supervised the study. All authors were involved in the acquisition, analysis, or interpretation of the data. SJS performed the statistical analyses. LM and SJS drafted the manuscript, and all authors critically revised it for important intellectual content. The corresponding author attests that all listed authors meet authorship criteria and that no others meeting the criteria have been omitted. SJS had full access to all the data in the study and are the guarantors.

## Funding

This study was supported by the UCSF Division of Hospital Medicine and the UCSF Health Office of Population Health.

## Sponsor’s Role

The funders had no role in study design, data collection and analysis, decision to publish, or preparation of the manuscript.

## Competing interests

All authors have completed the ICMJE uniform disclosure form at www.icmje.org/coi_disclosure.pdf (available on request from the corresponding author). LM, MG, LK, and SJS have nothing to disclose. KG reports that she was employed by the third party (CipherHealth) prior to her employment at UCSF and prior to any involvement with this study.

## Ethical approval

The Human Research Protection Program Institutional Review Board at the University of California, San Francisco, approved this study (IRB# 19-27987).

## Data sharing

Data used to complete this analysis used protected health elements. Researchers can contact the corresponding authors to request access to the study data. Code used to generate the cohort and perform the analyses can be found on github (https://github.com/sachinjshah).

## Appendix

### Appendix-1: Identification of Study Cohort

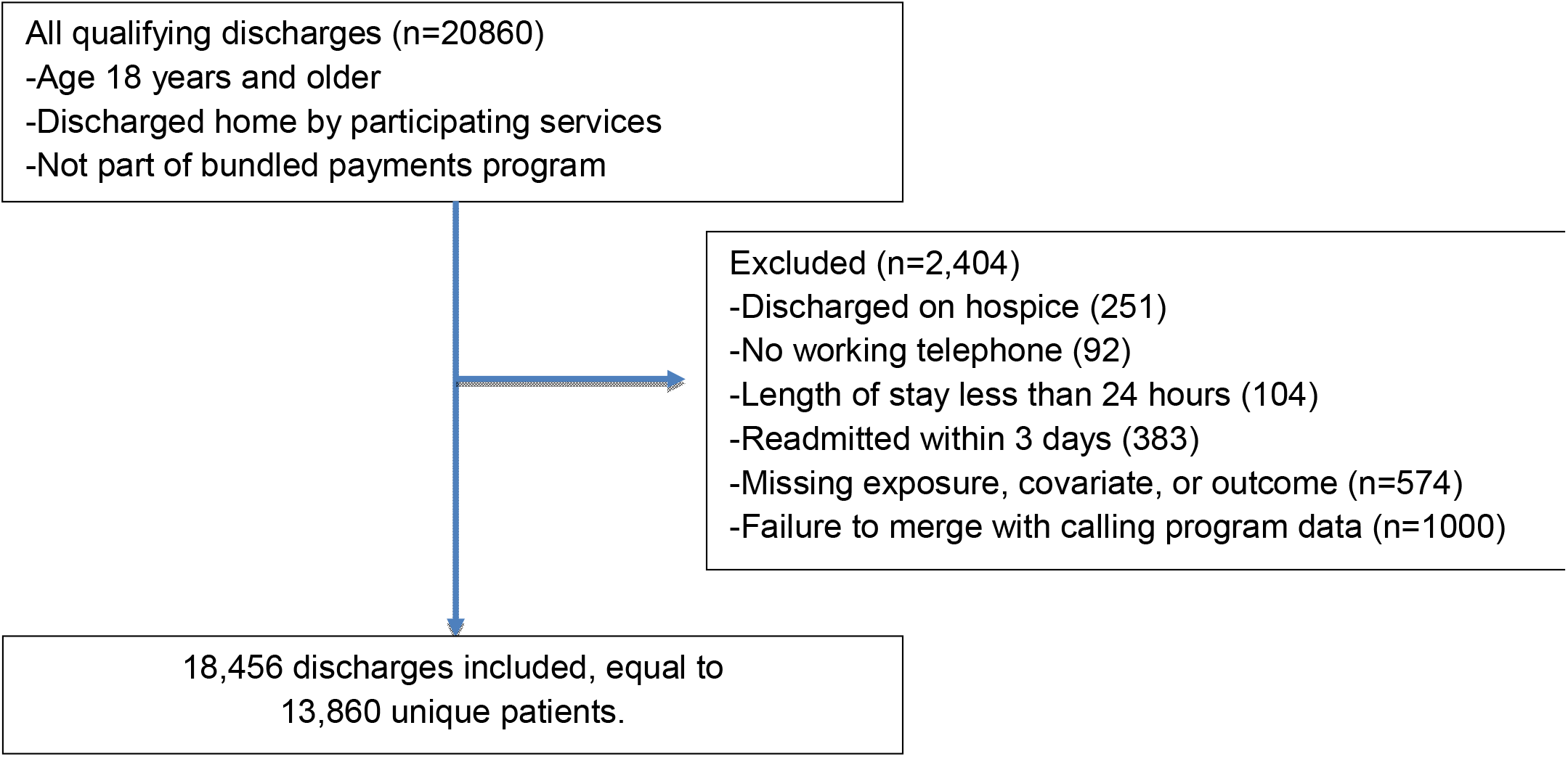

### Appendix-2: The Automated Script

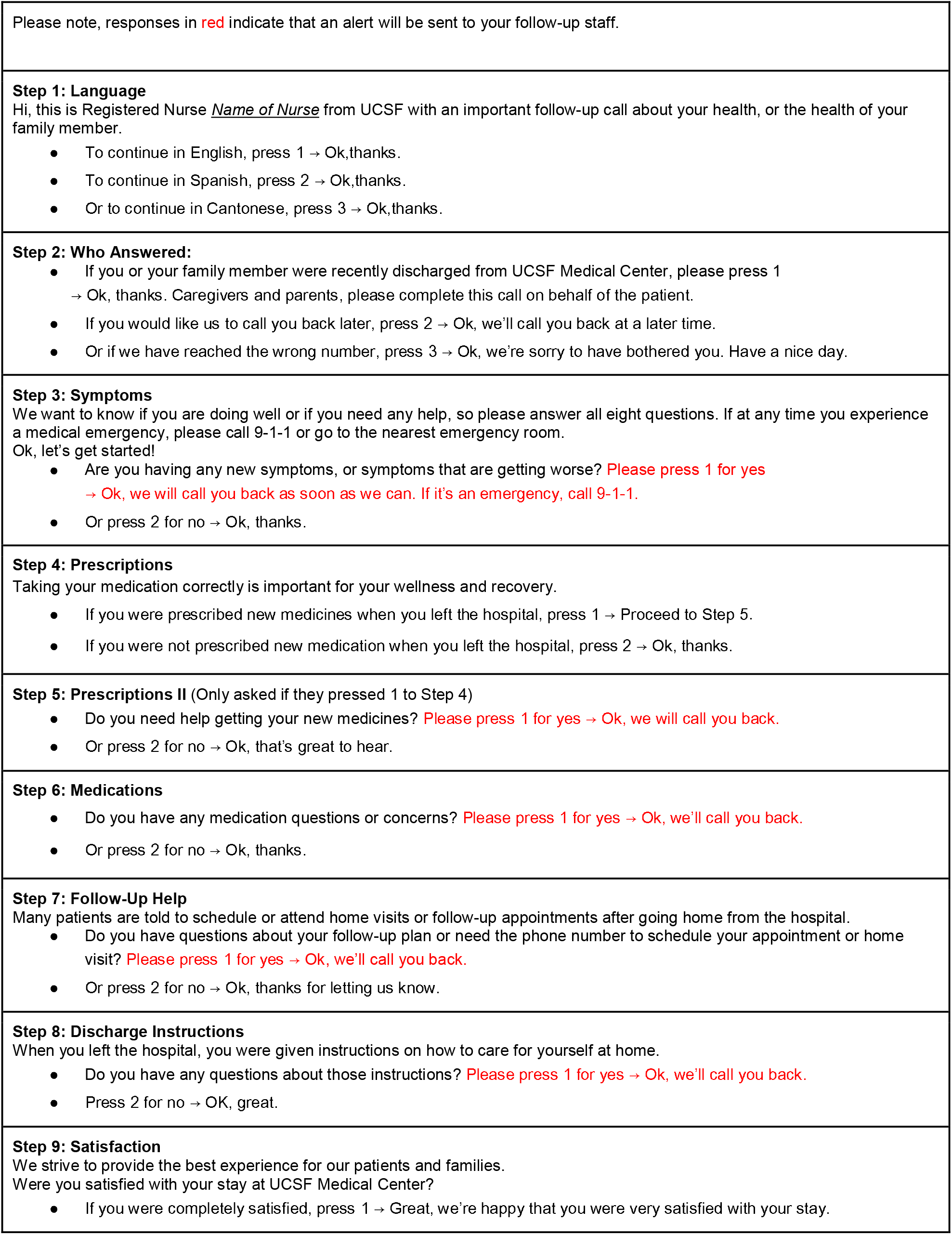
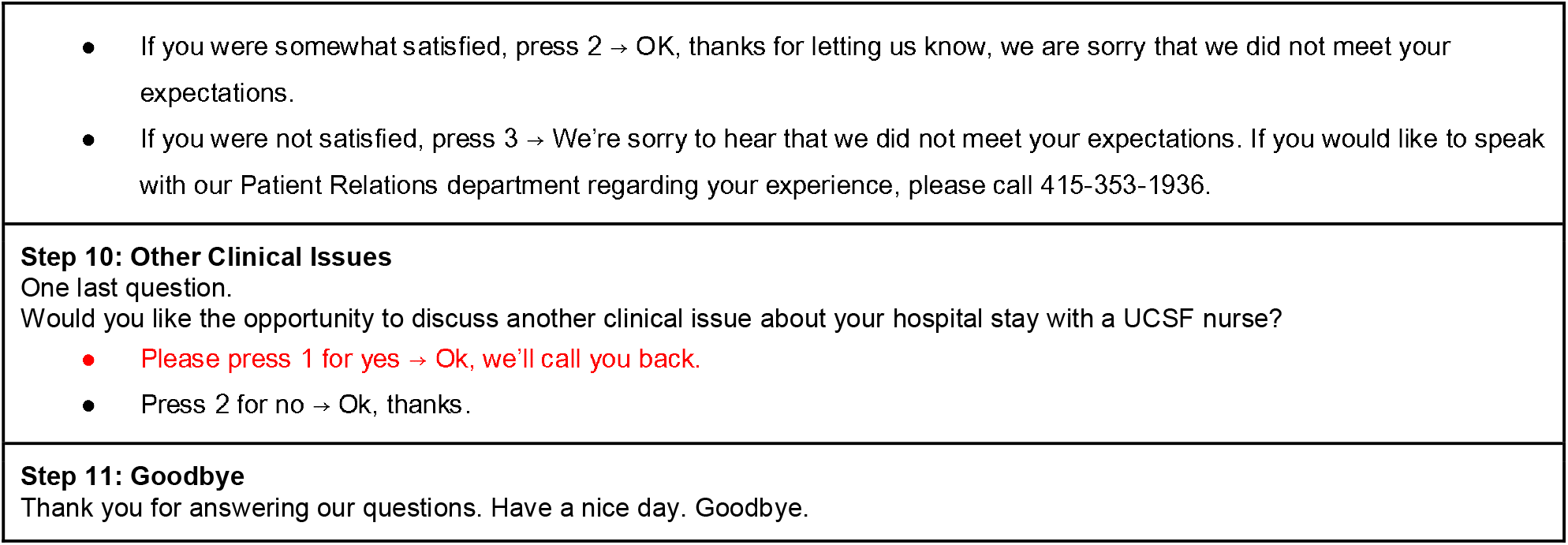

### Appendix-3: Unadjusted versions of main manuscript results

#### 3.1: Post-discharge program reach by English proficiency, unadjusted

Unadjusted results that correspond to Table 2 in the main manuscript

|  | Limited English<br>proficiency<br>(n=1566) | English<br>proficient<br>(n=12294) |
| --- | --- | --- |
| Answered automated call | 76.2% | 81.1% |
| Outreached completed by clinician | 8.1% | 4.6% |
| Reached by RN manual call | 5.7% | 0.7% |
| Not reached by any method | 10.1% | 13.7% |

#### 3.2 Patient-reported, post-discharge issues by English proficiency

Adjusted and unadjusted results that correspond to Figure 1 in the main manuscript

| Post-discharge issue | Unadjusted | Adjusted |
| --- | --- | --- |
| Discharge instructions |  |  |
| EP | 6.4% | 6.5% |
| LEP | 13.5% | 11.3% |
| Difference | 7.2% (95% CI, 5.1 to 9.2%) | 4.8% (95% CI, 2.7 to 6.9%) |
| Getting prescriptions |  |  |
| EP | 5.3% | 5.5% |
| LEP | 10.6% | 8.3% |
| Difference | 5.3% (95% CI, 3.1 to 7.6%) | 2.9% (95% CI, 0.6 to 5.1%) |
| Medication concerns |  |  |
| EP | 10.5% | 10.6% |
| LEP | 13.9% | 12.9% |
| Difference | 3.4% (95% CI, 1.3 to 5.5%) | 2.3% (95% CI, 0.0 to 4.6%) |
| Follow up questions |  |  |
| EP | 13.1% | 13.3% |
| LEP | 17.9% | 16.1% |
| Difference | 4.8% (95% CI, 2.5 to 7.2%) | 2.8% (95% CI, 0.3 to 5.3%) |
| New or worsening symptoms |  |  |
| EP | 11.9% | 11.9% |
| LEP | 14.8% | 15.1% |
| Difference | 2.8% (95% CI, 0.8 to 4.9%) | 3.2% (95% CI, 0.7 to 5.8%) |
| Any other clinical issues |  |  |
| EP | 12.6% | 13.0% |
| LEP | 20.4% | 16.6% |
| Difference | 7.8% (95% CI, 5.3 to 10.3%) | 3.6% (95% CI, 1.1 to 6.1%) |

#### 3.3 Time to issue resolution after discharge by English proficiency, unadjusted

Unadjusted results that correspond to Figure 2 in the main manuscript

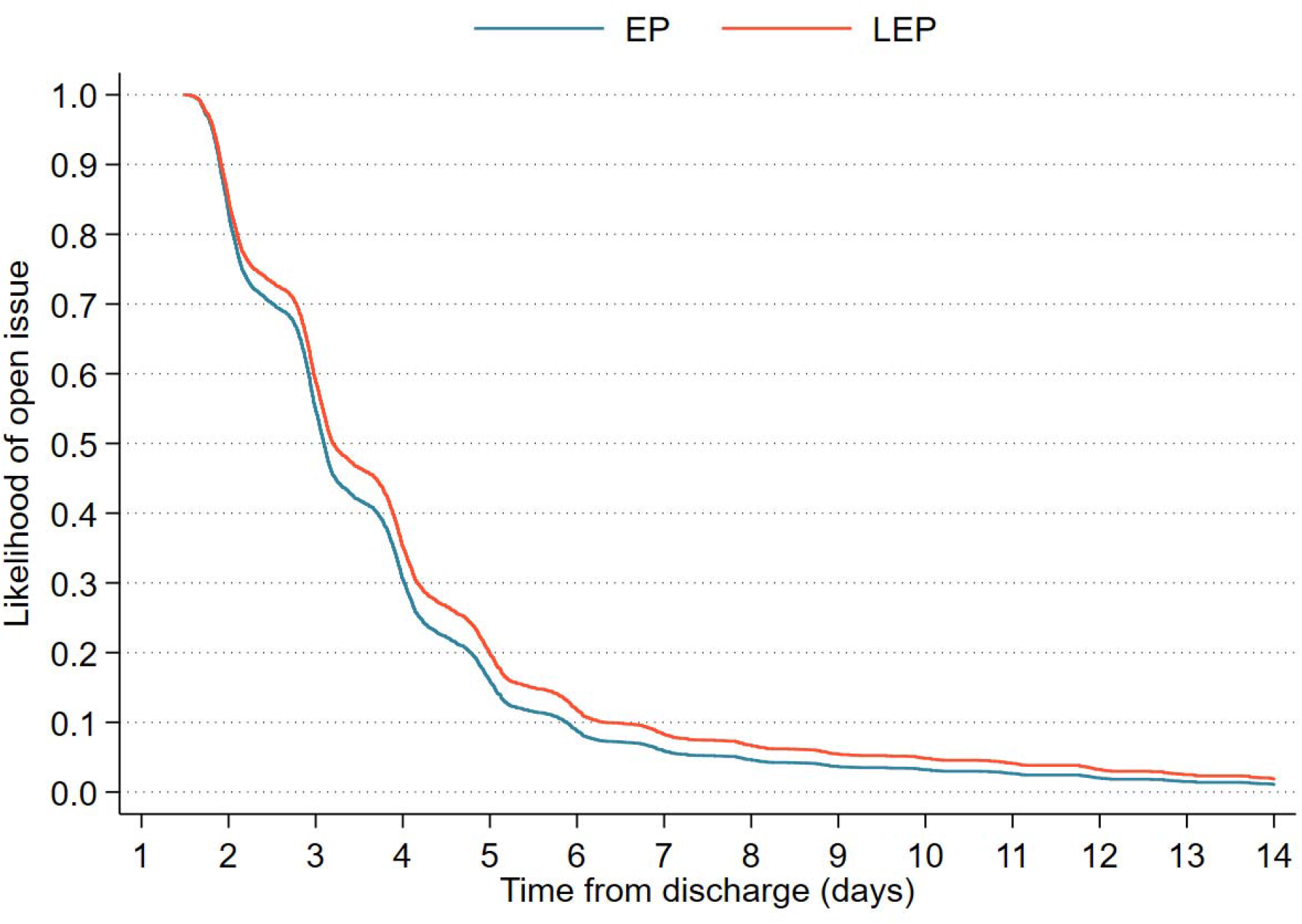

<u>Legend</u>

LEP - limited English proficiency, EP - English proficient

Plot depicts the likelihood of a patient’s reported issue being resolved at a given time post discharge by EP status. The likelihood is based on the Cox regression analysis that does not account for any confounders.

The unadjusted hazard of an open issue for patients with LEP is 0.88 (95%CI 0.80 to 0.96) relative to patients with EP. As presented in the main manuscript, Figure 2, the adjusted hazard of an open issue for patients with LEP is 0.84 (95%CI 0.75 to 0.93) relative to patients with EP.

### Appendix-4: Problem urgency as documented

When a problem was identified care transition nurses would document the problem in a note in the EMR with a structured header that denoted the urgency of the problem as they understood it. These categories include: RN spoke with the patient, clinician notified, clinician action requested, clinician action needed, not reached. These categories are hierarchical and mutually exclusive.

#### 4.1 Problem urgency by English proficiency, unadjusted

| Problem urgency by RN note | Limited English proficiency (n=555) | English proficient (n=3340) |
| --- | --- | --- |
| RN spoke with patient | 11.5% | 10.1% |
| Clinician notified | 59.1% | 55.2% |
| Clinician action requested | 11.4% | 15.4% |
| Clinician action needed | 1.6% | 1.6% |
| Not reached | 16.4% | 17.6% |

#### 4.2 Problem urgency by English proficiency, adjusted

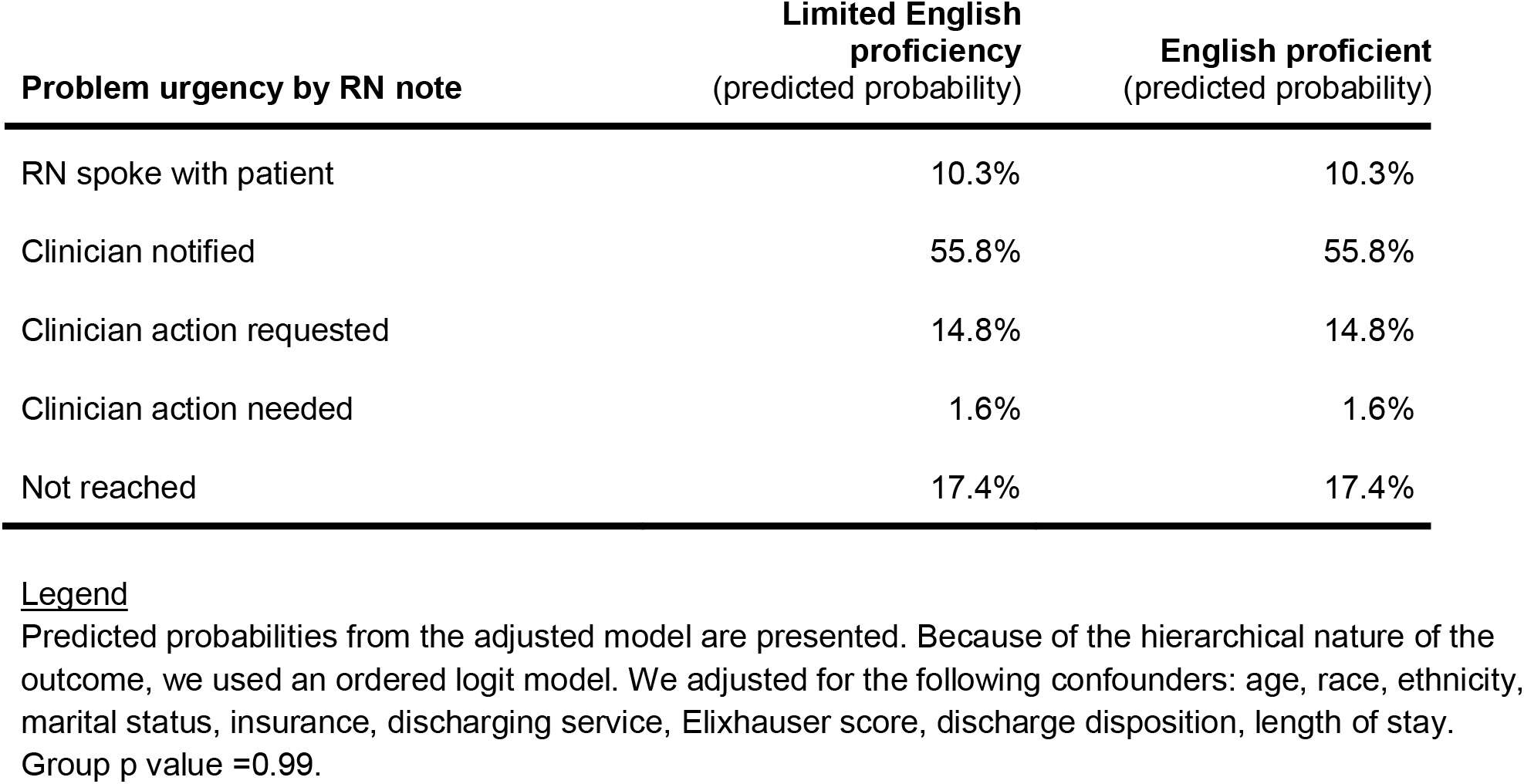

| Problem urgency by RN note | Limited English proficiency (predicted probability) | English proficient (predicted probability) |
| --- | --- | --- |
| RN spoke with patient | 10.3% | 10.3% |
| Clinician notified | 55.8% | 55.8% |
| Clinician action requested | 14.8% | 14.8% |
| Clinician action needed | 1.6% | 1.6% |
| Not reached | 17.4% | 17.4% |

### Appendix-5: Issue Escalation and Involvement of Other Professionals

|  | Unadjusted | Adjusted |
| --- | --- | --- |
| Escalation to nonclinical health staff |  |  |
| EP | 2.5% | 2.4% |
| LEP | 1.6% | 1.9% |
| Difference (LEP-EP) | -0.9% (-2.0 to 0.3%) | -0.6% (-2.1 to 0.9%) |
| Escalation to clinician |  |  |
| EP | 5.4% | 5.1% |
| LEP | 2.9% | 4.2% |
| Difference (LEP-EP) | -2.5% (-4.1 to -0.9%) | -1.0% (-3.4 to 1.4%) |
| Escalation to pharmacist |  |  |
| EP | 7.7% | 7.8% |
| LEP | 6.3% | 6.0% |
| Difference (LEP-EP) | -1.4% (-3.6 to 0.8%) | -1.8% (-4.2 to 0.6%) |

## Notes

### Funding Statement

This study was supported the UCSF Division of Hospital Medicine, and the UCSF Office of Population Health. The funders had no role in study design, data collection and analysis, decision to publish, or preparation of the manuscript.

### Summary of Updates

Revised cohort development figure and associated text.

